# SARS-CoV-2 Breakthrough Infections in Fully Vaccinated Individuals

**DOI:** 10.1101/2021.06.21.21258990

**Authors:** Esteban Ramirez, Rebecca P. Wilkes, Giovanna Carpi, Jack Dorman, Craig Bowen, Lisa Smith

## Abstract

**Importance:** While COVID-19 vaccines are highly effective against disease, breakthrough infections may occur in the context of rising variants of concern.

**Objective:** We paired random and passive surveillance nucleic acid testing with analysis of viral whole genomic sequences to detect and describe breakthrough infections, focusing in a university community.

**Design:** Anterior nasal swabs were collected from individuals for a nucleic acid amplification test (NAAT) for detection of SARS-CoV-2. A subset of NAAT positive samples was sequenced to determine variants associated with infections. Included in the testing and sequencing protocol were individuals that were fully vaccinated.

**Setting:** This study was performed as part of a surveillance program for SARS-CoV-2 on a university campus with 49,700 students and employees.

**Participants:** Surveillance testing was random and included approximately 10% of the population each week. Additionally, individuals self-identified with COVID-19 related symptoms or those that had close contact with SARS-CoV-2 positive individuals were also tested.

## Introduction

Severe acute respiratory syndrome coronavirus 2 (SARS-CoV-2) is the virus which causes COVID-19. Some infections are asymptomatic or cause mild symptoms, while other individuals are severely disabled or die from their disease. COVID-19 has caused 582,769 deaths in the United States to date. With the development and rapid rollout of COVID-19 vaccines, the positivity rate across the nation has started to decrease. Reinfections are reported from 2-12%^2,3^. A vaccine breakthrough infection is defined as the detection of SARS-CoV-2 RNA or antigen in a respiratory specimen collected from a person ≥14 days after they have completed all recommended doses of a U.S. Food and Drug Administration (FDA)-authorized COVID-19 vaccine (CDC), and reports of reinfections are accumulating^4^. Currently, in the United States, one is considered fully vaccinated two weeks after a two-dose vaccine (e.g., mRNA vaccines from Pfizer-BioNTech or Moderna) or two weeks after a one-dose vaccine (e.g., adenoviral vectored vaccine from Johnson & Johnson Janssen). At the time of this publication, variants of concern are on the rise. The most prevalent variant of concern within this population included here is B.1.1.7, followed distantly by P.1 and B.1.429. Understanding these breakthrough infections are important as it can help guide public health officials, pharmaceutical industries, and medical providers to determine the next steps in development of booster vaccines, efficacy of our vaccines against reinfection and find out if the current vaccines are limiting infections if exposed to variants.

## Methods

This is a retrospective analysis of results that were obtained from students and employees in a university setting. Data were obtained in deidentified format from the electronic health record system from One-to-One Health. Approval from the One-to-One Health review board was obtained. The Institutional Review Board from the Purdue University Human Research Protection Program determined that viral genome sequencing of deidentified remnant RNA samples included in this study is not research involving human subjects (IRB-2021-438). Individuals were tested either through a random surveillance program or because of COVID-19-like symptoms or exposure. Due to the testing protocol at the time of this publication, individuals that were fully vaccinated were still included in surveillance testing. Anterior nasal swabs were collected in PrimeStore MTM (Longhorn Vaccines and Diagnostics) and submitted to the Animal Disease Diagnostic Lab at Purdue University for a nucleic acid amplification test (NAAT) (TaqPath COVID-19 Test Kit, ThermoFisher Scientific). Viral whole genome sequencing was performed on at least 15% (34% on average) of positive samples per week in the Carpi Lab using MinION Mk1B or GridION Mk1 Nanopore devices (Oxford Nanopore Technologies, ONT, UK). High-accuracy basecalling was performed using Guppy v4.2.4 and consensus genome sequences were generated following the ARTIC bioinformatics pipeline v.1.1.3. Lineages were assigned using Pangolin v.2.4.2, according to the system described in Rambaut et al 2020^5^. Samples that were sequenced were randomly selected based on Ct values (Cts ≤30 could be included). Additionally, samples were specifically selected for sequencing, including S drop out samples (no spike gene detected but other two targets were positive), samples from those with history of travel (both within and outside the US), and samples from fully vaccinated individuals and from those with reinfections. This report reviews data from 14 individuals that were fully vaccinated.

## Results

The data from our cohort demonstrated that from February 10, 2021 to May 10, 2021, we performed 2,551 nucleic acid amplification tests from samples from fully vaccinated individuals, and of these, 14 represent breakthrough infections (0.55%, 95% CI: 0.30 to 0.92). In comparison, we tested 65,877 samples from unvaccinated/partially vaccinated individuals during the same time period, of which 1,482 were positive by NAAT (2.25%, 95% CI: 2.14 to 2.37). Of the breakthrough infections, six of these individuals received the Pfizer vaccine, five received the Moderna vaccine, and three received the J&J vaccine. Some of these individuals carried high viral loads, based on Ct values (Table 1). Many of these breakthroughs were associated with variants of concern (VOCs -B.1.1.7 and P.1) according to the CDC classification (Table 1). Asymptomatic infections composed 9/14 (64%) of the cases, and the majority of cases were detected in females (10/14, 71%).

**Table 1.** SARS-CoV-2 Breakthrough infections after fully vaccinated status.

| **Gender** | **Age** | **Vaccine** | **Symptomatic** | **Ct values\*** | **Virus lineage<sup>#</sup>** | **GISAID Accession ID** |
| --- | --- | --- | --- | --- | --- | --- |
| Female | 16-25 | Pfizer | no | 33.56,<br>34.36,<br>33.16 | N/A | N/A |
| Female | 16-25 | Pfizer | no | 32.44,<br>31.36,<br>32.44 | N/A | N/A |
| Male | 16-25 | Pfizer | no | 33.67,<br>36.01,<br>ND | N/A | N/A |
| Female | 16-25 | Moderna | no | 34.77,<br>34.1,<br>34.79 | N/A | N/A |
| Female | 16-25 | Moderna | no | 33.67,<br>36.92,<br>35.01 | N/A | N/A |
| Female | 46-55 | Moderna | no | 22.43,<br>22.81,<br>ND | B.1.1.7 | EPI_ISL_1823539 |
| Female | 16-25 | Moderna | yes | 22.58,<br>21.73,<br>ND | B.1.1.7 | EPI_ISL_2304374 |
| Female | 16-25 | Pfizer | no | 14.48,<br>14.35,<br>ND | B.1.1.7 | EPI_ISL_2304375 |
| Female | 16-25 | Moderna | yes | 28.04,<br>27.55,<br>ND | B.1.1.7 | EPI_ISL_2304382 |
| Male | 16-25 | J&J | no | 25.83,<br>26.55,<br>25.91 | P.1 | EPI_ISL_2304422 |
| Female | 16-25 | J&J | yes | 15.14,<br>15.56,<br>15.7 | P.1 | EPI_ISL_2304452 |
| Male | 36-45 | Pfizer | no | 23.19,<br>23.43,<br>24.51 | B.1.2 | EPI_ISL_2304450 |
| Male | 16-25 | Pfizer | yes | 15.55,<br>17.26,<br>14.14 | B.1.526.2 | EPI_ISL_1823443 |
| Female | 56-65 | J&J | yes | 16.39,<br>17.69,<br>16.85 | B.1.617.2 | EPI_ISL_2304456 |

\*Ct values are provided for viral targets ORF1ab, N, and S, respectively. ND (not detected).

Contact tracing reports for these 14 individuals were reviewed to assess if their identified close contacts tested positive afterward. One of 13 cases (one report was unavailable for review) had a close contact test positive. This one positive was caused by a B.1.1.7 VOC.

## Discussion

B.1.1.7 has been shown in multiple studies to have minimal impact on neutralization by post-vaccination sera^6^. It is probable that the breakthroughs with this particular variant are more likely a result of increased circulation of this variant during this time point. This variant composed approximately 31% of the total positive samples during the period of this particular breakthrough, with at least 12 other variants composing the other 69% in this population. P.1 contains the E484K mutation, which mediates escape from vaccine-induced humoral immunity^7^. Notable mutations in the breakthrough infection associated with the B.1.617.2 lineage includes L452R and P681R in the spike protein. P681R mutation is found in the furin cleavage site, which mediate increased rate of membrane fusion, internalization, leading to higher viral loads and increased transmissibility^8^. Interestingly, B.1.2 has markedly declined within this population, so detection of this lineage in association with one vaccine breakthrough case was an interesting finding, however no notable mutations except for the linked D614G and P314L were detected in this B.1.2 viral genome. These data demonstrate that each of the vaccines have associated breakthrough infections, and with these small numbers we cannot report that one is more or less effective in our population. Of note, the J&J vaccine during this period was temporarily removed from the market due to investigation. Females have composed approximately 63% of reported breakthrough cases^9^. Females are overrepresented in this group as well. While approximately 45% of vaccine breakthroughs have been reported from those aged 60 and above, these data show that these infections also occur in college-aged individuals. Given that the majority of these breakthroughs were detected through surveillance from asymptomatic individuals on a university campus, it is likely that this age group and asymptomatic infections are underrepresented with regard to reported vaccine breakthrough cases. However, the data reviewed in this report suggests that these breakthrough infections may not have much clinical significance because only 1 in contact case was identified. However, it is important to note that once identified as positive for SARS-CoV-2, these individuals were placed in isolation so further spread would be minimized or prevented.

## Conclusion

These results provide genomic evidence of breakthrough infections and suggest that fully vaccinated individuals, including those with asymptomatic infections, are less likely to serve as a source of infection for others; however, this finding requires further investigations. Notably, the percentage of positives in this group was significantly lower than in the unvaccinated/partially vaccinated cohort. Fewer detected breakthrough infections in the full-vaccinated group reinforces the need to get vaccinated in order to decrease the spread of SARS-CoV-2 infection. Our observations provide support for the need of sustained efforts to diagnose asymptomatic infections and characterization of variants.

## Data Availability

Genomic data are available on GISAID (see Table 1 for accession numbers).

https://www.gisaid.org

